# Identifying therapeutic targets for cancer: 2,094 circulating proteins and risk of nine cancers

**DOI:** 10.1101/2023.05.05.23289547

**Authors:** Karl Smith-Byrne, Åsa Hedman, Marios Dimitriou, Trishna Desai, Alexandr V. Sokolov, Helgi B. Schioth, Mine Koprulu, Maik Pietzner, Claudia Langenberg, Joshua Atkins, Ricardo Cortez, James McKay, Paul Brennan, Sirui Zhou, Brent J. Richards, James Yarmolinsky, Richard M. Martin, Joana Borlido, Xinmeng J. Mu, Adam Butterworth, Xia Shen, Jim Wilson, Themistocles L. Assimes, Rayjean J. Hung, Christopher Amos, Mark Purdue, Nathaniel Rothman, Stephen Chanock, Ruth C. Travis, Mattias Johansson, Anders Mälarstig

**Affiliations:** Cancer Epidemiology Unit, Oxford Population Health, University of Oxford, Oxford, United Kingdom; External Science and Innovation, Pfizer Worldwide Research, Development and Medical, Stockholm, Sweden; Department of Medicine, Department of Medicine, Stockholm, Sweden; Department of Surgical Sciences, Functional Pharmacology and Neuroscience, Uppsala University, Uppsala, Sweden; MRC Epidemiology Unit, University of Cambridge, Cambridge, United Kingdom; Computational Medicine, Berlin Institute of Health at Charité – Universitätsmedizin Berlin, Berlin, Germany; Precision Healthcare Institute, Queen Mary University of London, London, United Kingdom; Genomic Epidemiology Branch, International Agency for Research on Cancer (IARC-WHO), Lyon, France; Department of Human Genetics, McGill University, Montréal, Quebec, Canada; Departments of Medicine (Endocrinology), Human Genetics, Epidemiology and Biostatistics, McGill University, Montréal, Quebec, Canada; MRC Integrative Epidemiology Unit, University of Bristol, Bristol, United Kingdom; Population Health Sciences, Bristol Medical School, University of Bristol, Bristol, United Kingdom; NIHR Bristol Biomedical Research Centre, Hospitals Bristol and Weston NHS Foundation Trust and the University of Bristol, Bristol, United Kingdom; Cancer Immunology Discovery, Pfizer Worldwide Research and Development Medicine, Pfizer Inc., San Diego, United States of America; Oncology Research Unit, Pfizer Worldwide Research and Development Medicine, Pfizer Inc, San Diego, United States of America; Department of Public Health and Primary Care, University of Cambridge, Cambridge, United Kingdom; Usher Institute, MRC Human Genetics Unit, University of Edinburgh, Edinburgh, United Kingdom; Division of Cardiovascular Medicine and the Cardiovascular Institute, School of Medicine, Stanford University, Stanford, United States of America; Prosserman Centre for Health Research, Lunenfeld-Tanenbaum Research Institute, Sinai Health System and University of Toronto, Toronto, Canada; Department of Medicine, Epidemiology Section, Institute for Clinical and Translational Research, Baylor Medical College, Houston, United States of America; Division of Cancer Epidemiology and Genetics, National Cancer Institute, Rockville, United States of America; Occupational and Environmental Epidemiology Branch, Division of Cancer Epidemiology and Genetics, National Cancer Institute, Rockville, United States of America

**Keywords:** Proteomics, Cancer, cis pQTL

## Abstract

**Background:** Understanding the role of circulating proteins in cancer risk can reveal key biological pathways and identify novel therapeutic targets for cancer prevention.

**Methods:** We investigated the associations of 2,094 circulating proteins with risk of nine common cancers (bladder, breast, endometrium, head and neck, lung, ovary, pancreas, kidney, and malignant non-melanoma) using *cis* pQTL Mendelian randomisation (MR) and colocalization. Findings for proteins with support from both MR, after correction for multiple-testing, and colocalization were replicated using an independent cancer GWAS. Additionally, MR and colocalization phenome-wide association analyses (PHEWAS) were conducted to identify potential adverse side-effects of altering risk proteins. Finally, we mapped cancer risk proteins to drug and ongoing clinical trials targets.

**Results:** We identified 40 proteins associated with cancer risk, of which a majority replicated and were novel. Among these were proteins associated with common cancers, such as PLAUR and risk of breast cancer [odds ratio per standard deviation increment (OR): 2.27, 95% CI: 1.88 to 2.74], and with high-mortality cancers, such as CTRB1 and pancreatic cancer [OR: 0.79, 95% CI: 0.73 to 0.85]. PHEWAS highlighted multiple links between proteins and potential adverse effects of protein-altering interventions. Additionally, 18 proteins associated with cancer risk mapped to existing therapeutic interventions, while 15 were not currently known to be under clinical investigation, such as GAS1 and triple negative breast cancer [OR: 1.88, 95% CI: 1.42 to 2.47].

**Conclusion:** Our findings emphasize the importance of proteomics for improving our understanding of cancer aetiology. Additionally, we demonstrate the benefit of in-depth protein PHEWAS analyses on risk proteins to identify potential adverse side-effects of protein-altering interventions. Using these methods, we identify a subset of risk proteins as potential drug targets for the prevention and treatment of cancer as well as opportunities for drug repurposing.

## Introduction

Proteins govern cellular action in all human biological processes and are crucial for our defences against both the onset and progression of cancer. Identifying novel circulating proteins important to the aetiology of cancer may improve our understanding of pathways leading to cancer and highlight potential targets for therapeutic prevention. Circulating proteins are valuable candidate targets for drug development since drug-target engagement can be evaluated in the bloodstream during randomised control trials (RCTs), accelerating target development. Additionally, identifying circulating cancer risk proteins allows for the subsequent selection of future RCT participants with risk-inducing protein concentrations, which may improve RCT effectiveness. Developing therapeutic prevention strategies, either alone or as a complement to existing prevention programs, such as smoking cessation, are urgently needed given cancer burden is projected to double by the year 2040^1^.

Therapeutic prevention is an effective and commonly used strategy for the primary prevention of some common chronic diseases. Prevention strategies have thus far been most successful for cardiovascular disease using statins that target the HMG-CoA reductase protein as a first line treatment to lower low-density lipoprotein (LDL) cholesterol^2,3^. In contrast, efforts to identify targets for the therapeutic prevention of cancer have been less fruitful, hampered by a more complex aetiology and difficulty identifying potential targetable aetiological biomarkers^4^. Exceptions include therapeutically targeting the oestrogen receptor (ER) to prevent breast cancer^5^ and COX2 to prevent colorectal cancer in high-risk individuals^6^. Additional aetiological proteins for specific cancers have been identified, such as the role of higher levels of insulin-like growth factor-I in the development of breast^7^, colorectal^8^, and prostate^9,10^ cancers, and of higher microseminoprotein-beta with lower prostate cancer risk ^11^. Together these examples highlight the opportunity that may result if aetiological proteins for cancer are identified and the feasibility of using these to develop novel therapeutic prevention tools where high-risk populations are well-defined.

Identifying candidate aetiological biomarkers for cancer risk has traditionally involved analysing specific hypothesis-driven markers for single cancer outcomes in pre-diagnostic samples and comparable controls from large prospective cohorts^9,12,13^. The advent of high-throughput platforms that can measure hundreds to thousands of biomarkers simultaneously using small sample volumes has enabled hypothesis-free discovery analyses, but costs remain prohibitively high. An alternative cost-effective approach, that also limits bias by confounding and reverse causation, is to use robust genetic proxies of blood biomarkers to evaluate their aetiological relevance along the lines of Mendelian randomisation (MR)^14,15^. Using such MR-based approaches facilitates simultaneously querying thousands of markers in relation to risk of multiple cancers using genome-wide association data, which can identify novel risk markers and assess their association with one or multiple cancers. Proteins represent a particularly appealing application of MR as the blood concentrations of many proteins are regulated by genetic variants, many of which lie in or near a protein’s cognate gene (variants known as *cis* protein quantitative trait loci [*cis* pQTL])^16^. *Cis* pQTL likely influence biological processes directly, such as by transcription or translation, making them less prone to common sources of bias in MR studies like horizontal pleiotropy^17^. It is also possible to complement *cis* pQTL-based MR analyses with colocalization analyses to further exclude confounding by linkage disequilibrium^18^. These methodologies allow for the *in-silico* simultaneous evaluation the role of thousands of proteins in the aetiology of common cancers with high specificity.

In the current study, we estimated the associations of 2,094 circulating proteins with risk of nine common cancers using data from a total of 337,822 cancer cases. We aimed to identify novel cancer risk proteins and to assess whether these proteins may cause multiple or specific cancers. Where possible, we mapped risk proteins to potential therapeutic interventions and used MR and colocalization phenome-wide association analyses (PHEWAS) to describe the promise and complexities that may result from intervening on risk proteins in terms of potential adverse outcomes.

## Methods

### Overall Study Design

We sought to identify novel aetiological proteins for nine common cancers, including cancer of the head and neck, lung, kidney, pancreas, bladder, breast, ovary, and endometrium, as well as malignant non-melanoma. We used MR to evaluate the association of 2,094 blood protein concentrations with cancer risk based on *cis* pQTL single nucleotide polymorphisms (SNPs). We subsequently performed colocalization analyses for loci where MR indicated a nominally significant association with cancer risk, to assess the presence of confounding by linkage disequilibrium (LD). Where other independent sources of cancer GWAS were available, we also performed an external validation of *cis* pQTL associations with cancer risk. We then conducted an MR and colocalization PHEWAS as well as a review of public databases to assess whether proteins identified in our analyses as cancer risk factors were also associated with other important characteristics or diseases, which may inform potential adverse effects of future protein-altering interventions. Finally, we mapped cancer risk proteins to targets for approved drugs and those being evaluated in ongoing clinical trials.

### Protein effects on cancer risk

#### Collection and quality control for cis pQTL

We gathered summary statistics from publicly available protein GWAS’^15,19–21^ and extracted all independent *cis* pQTL [clumping at r^2^ < 0.01], defined as independent SNPs associated with a protein concentration in blood lying within 1 megabase of a protein’s cognate gene with at least p < 5e-08. We additionally re-processed previously published protein GWAS on the OpenGWAS platform to identify additional *cis* pQTL significant at p < 5e-05 due to strong *a priori* for SNP associations with protein concentrations at or near by its cognate gene (See Supplementary Table 1 and Supplementary Methods). *Cancer GWAS summary statistics*. Nine cancer outcomes and their subtypes (where applicable/available) were considered in this study, including cancer of the bladder, breast, endometrium, head and neck, lung, ovary, pancreas, kidney, and malignant non-melanoma (Table 1. and Supplementary methods).

**Table 1.** Description of cancer risk GWASs including ICD-10 codes, case and control counts, and study reference.

| Cancer | ICD-10 | Case/Control | Ref |
| --- | --- | --- | --- |
| Bladder | C67 | 8,988/11,978 | 22 |
| Breast* | C50 | 133,384/113,789 | 23 |
| Endometrium | C54.1 | 12,906/108,979 | 24 |
| Head and neck* | C02.0–C02.9, C03.0–C03.9, C04.0–C04.9, C05.0–C06.9, C01.9, C02.4, C09.0–C10.9 | 6,034/6,585 | 25 |
| Lung*,** | C34 | 41,477/105,297** | 26 |
| Ovary* | C56 | 25,509/40,941 | 27 |
| Pancreas | C25 | 7,638/7,364 | 28 |
| Kidney | C64 | 10,784/20,406 | 29 |
| Malignant non-melanoma | C43 | 23,694/372,016 | 30 |

\* Cancer subtypes included and described in supplementary methods.
\*\* Total effective sample size from a meta-analysis between GWAS of family history and GWAS in INTEGRAL-ILCCO.

#### MR and colocalization analyses

*Cis* pQTL were harmonised with GWAS summary statistics for each cancer outcome by matching on rsID where directly available, and by selecting a proxy where necessary. Primary risk estimates were odds ratios estimated using per-*cis* pQTL Wald Ratios. All MR associations with *p*_Wald_ < 0.05 were subsequently evaluated for confounding by LD and the probability of a shared causal locus (PP4) between protein concentrations and cancer risk using two approaches: conventional colocalization^18,31^ and conditional iterative colocalization^18^. The greatest PP4 from these two colocalization methods [PP4_max_] was used and we considered PP4_max_ > 0.7 as indicating *cis* pQTL MR associations were unlikely to have be confounded by LD. *Cis* pQTL with Bonferroni significant associations (i.e., *p*_Wald_ < 0.05/N_Proteins_ where N_Proteins_ is the number of unique proteins analysed for a given cancer outcome) that also had evidence of colocalization (i.e., PP4_max_ > 0.7) were subjected to further follow-up analyses. Further details in supplementary methods.

#### Replication of candidate aetiological proteins for cancer risk

Where data were available, we conducted a replication of noteworthy *cis* pQTL MR associations (i.e. PP4_max_ > 0.7 & Bonferroni *p*_Wald_) using external cancer GWAS data from either a meta-analysis of FinnGen r9^32^ and the UK Biobank^33^, or from FinnGen alone depending on the endpoint (see Table 2 for case counts and Supplementary Methods).

**Table 2.**
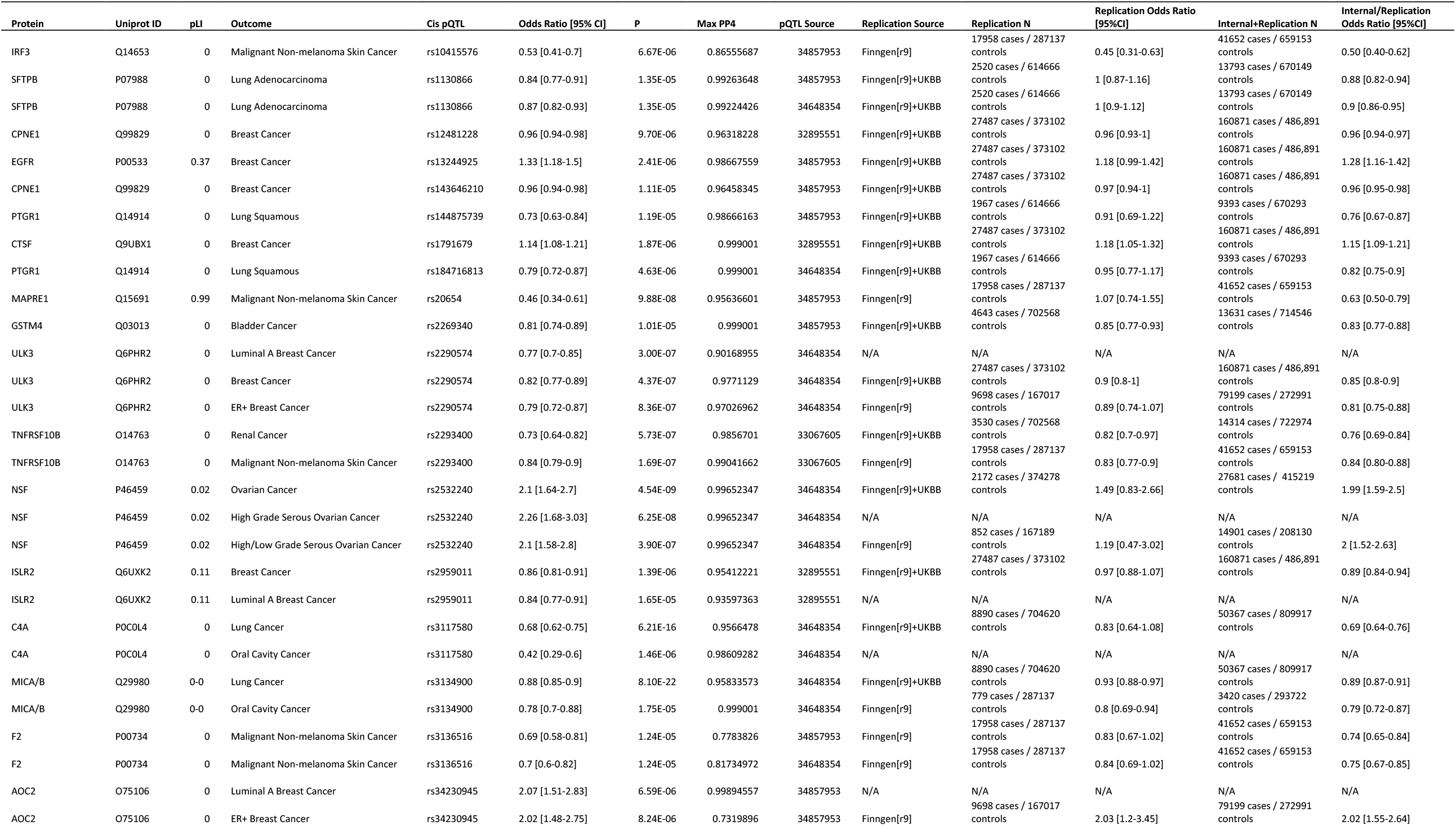
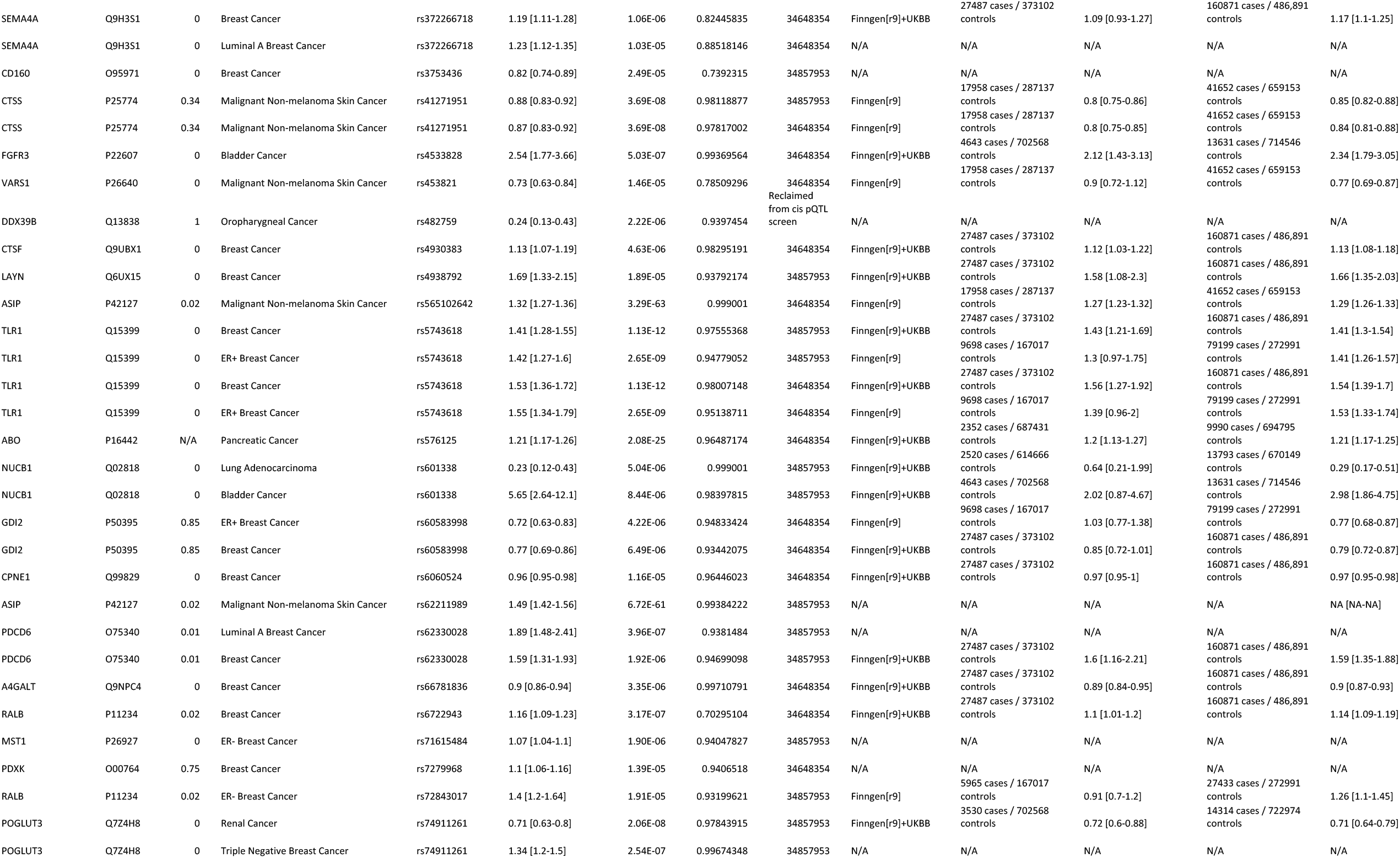
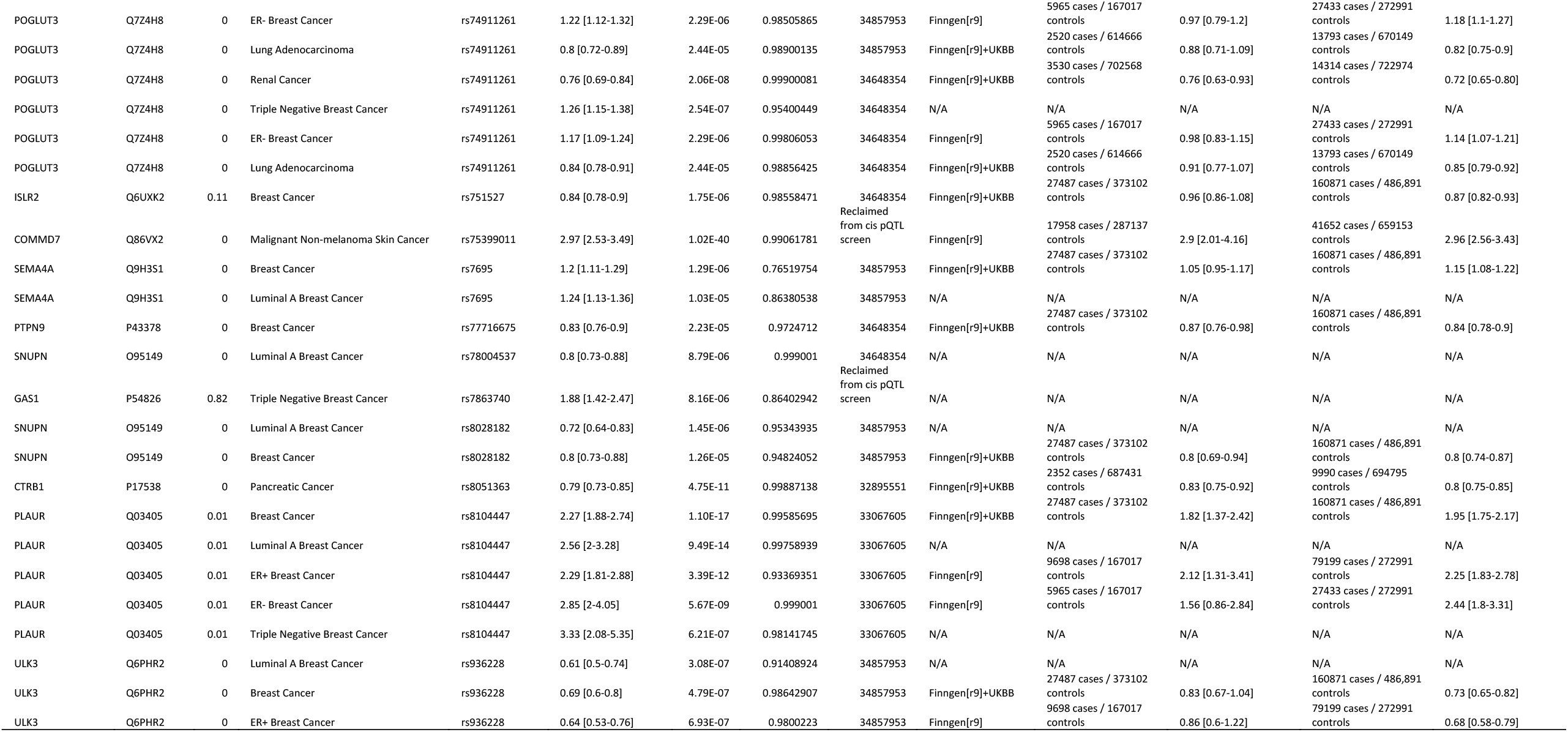
Main results for the association of genetically predicted protein levels with risk of cancer including Mendelian randomisation and colocalization analyses and external replication analyses, where available.

### Protein associations with other traits

We conducted additional analyses to provide greater context to the specificity of an identified cancer risk protein association using PHEWAS MR and colocalization analyses as well as consulting several public databases. We performed these steps to collate information on potential harmful or additional beneficial consequences of altering identified protein concentrations in human populations. Firstly, we assessed the association of each cancer risk protein with all available traits on the OpenGWAS platform using MR and colocalization methods as previously described^34^. We additionally used HyprColoc^35^ to assess whether any subset of protein-trait associations we identified may elucidate a potential causal pathway between risk of cancer and the indicated risk protein. Secondly, we conducted a search of several relevant databases that included information on probability of loss of function intolerance (pLI), exome-sequencing studies, rare-variant association studies, and Mendelian genetics not likely to overlap with OpenGWAS traits. Further details in supplementary methods.

### Drug Target pQTL Analyses

A secondary analysis was conducted restricting MR and colocalization analyses to *cis* pQTL with a cognate gene that is an established pharmaceutical target for the action of one of 867 existing drugs identified by reference to a combination of databases (including Drugbank and ClinicalTrials.gov) and expert curation (Supplementary methods). We defined noteworthy drug target proteins as having *p*_Wald_ < 0.05/N_Proteins_ where N_Proteins_ is the number of unique proteins analysed for a given cancer outcome that were identified as the cognate gene of a pharmaceutical target. We additionally queried the Cortellis database (https://www.cortellis.com/ Clarivate Analytics) to assess the highest current level of clinical development stage for proteins identified to associate with cancer risk from main analyses.

All analysis was performed using R version 4.1.2.

## Results

### Protein Effects on Cancer Risk

In total, 4,874 of the 5,117 *cis* pQTL were available for analysis with at least one cancer site [min: 3,508 *cis* pQTL for bladder cancer and max: 4,576 *cis* pQTL for kidney cancer], which represented 2,093 of 2,175 proteins with *cis* pQTL included in our study [min: 1,723 for bladder cancer and max: 1,997 proteins for skin cancer] (Fig 1.). MR and colocalization analyses identified 40 proteins (Table 2, Fig 2.) with an association with at least one cancer site [min: one protein for ovarian cancer, max: 21 proteins for breast cancer]. A further 433 proteins were identified to have evidence of colocalization [PP4 > 0.7] and at least a nominally significant MR association with risk of cancer [min: 8 proteins for malignant non-melanoma and max: 242 for Breast Cancer] (Figure 1, Supplementary Table S2). We observed limited evidence for the association of proteins with risk for clear cell ovarian cancer, ever smoking lung cancers, HER2 enriched, luminal B, and luminal B-HER2 negative breast cancers. Additionally, we did not identify any proteins as a risk factor for cancer using multiple, independent *cis* pQTL in MR analyses. Results by cancer site are summarised below.

**Figure 1.**
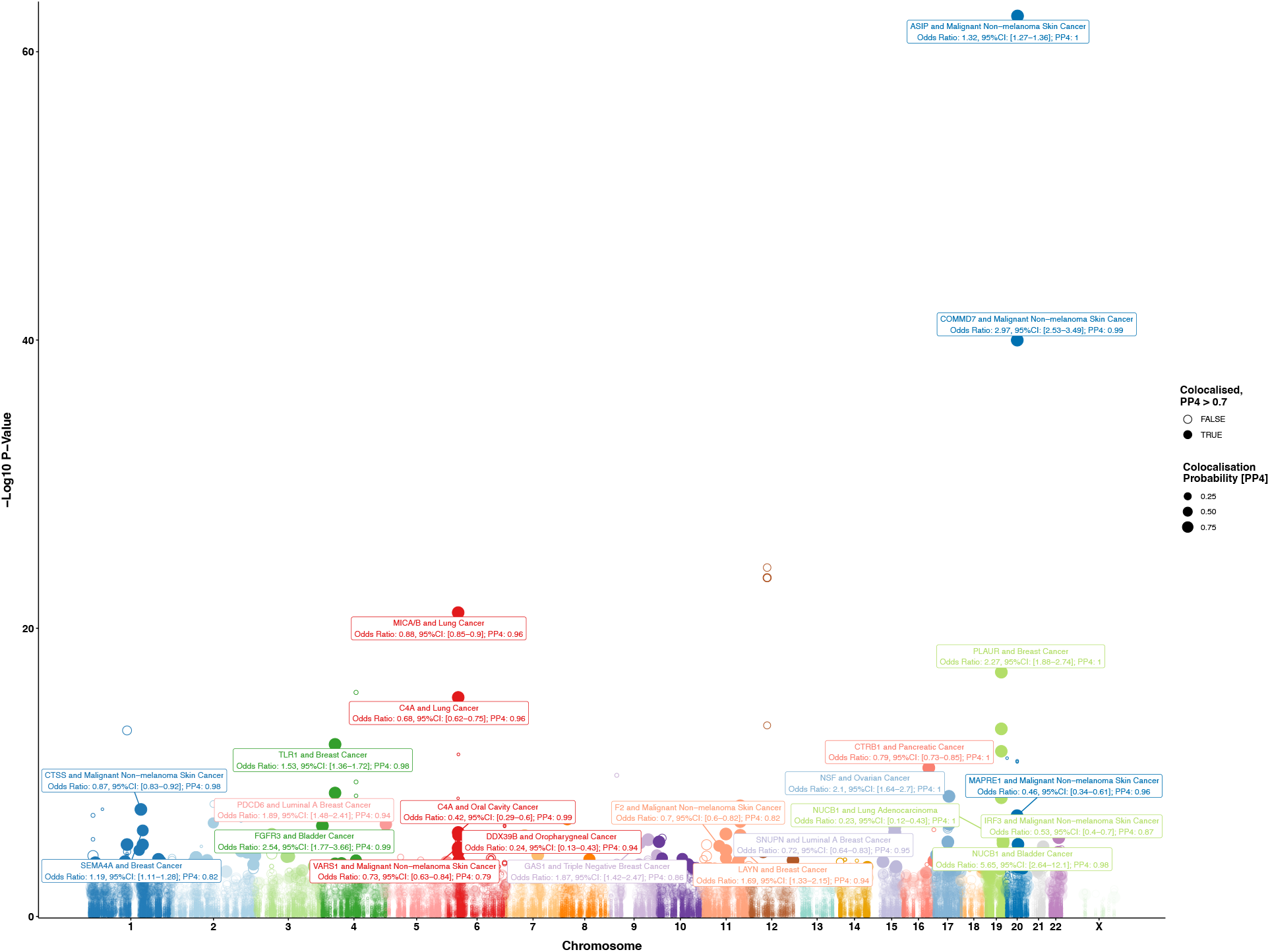
Association of genetically predicted protein concentrations with cancer risk presented as a Manhattan plot where position is given by *cis* pQTL coordinate with a selection of cancer risk associations additionally labelled for their association with cancer risk and colocalization probability (PP4). Points highlighted as filled-in are those with PP4 > 0.7 with point size reflecting PP4 magnitude, which can vary between 0 and 1. Risk associations with MR *p* > 0.05 were not subject to colocalization analyses.

**Figure 2.**
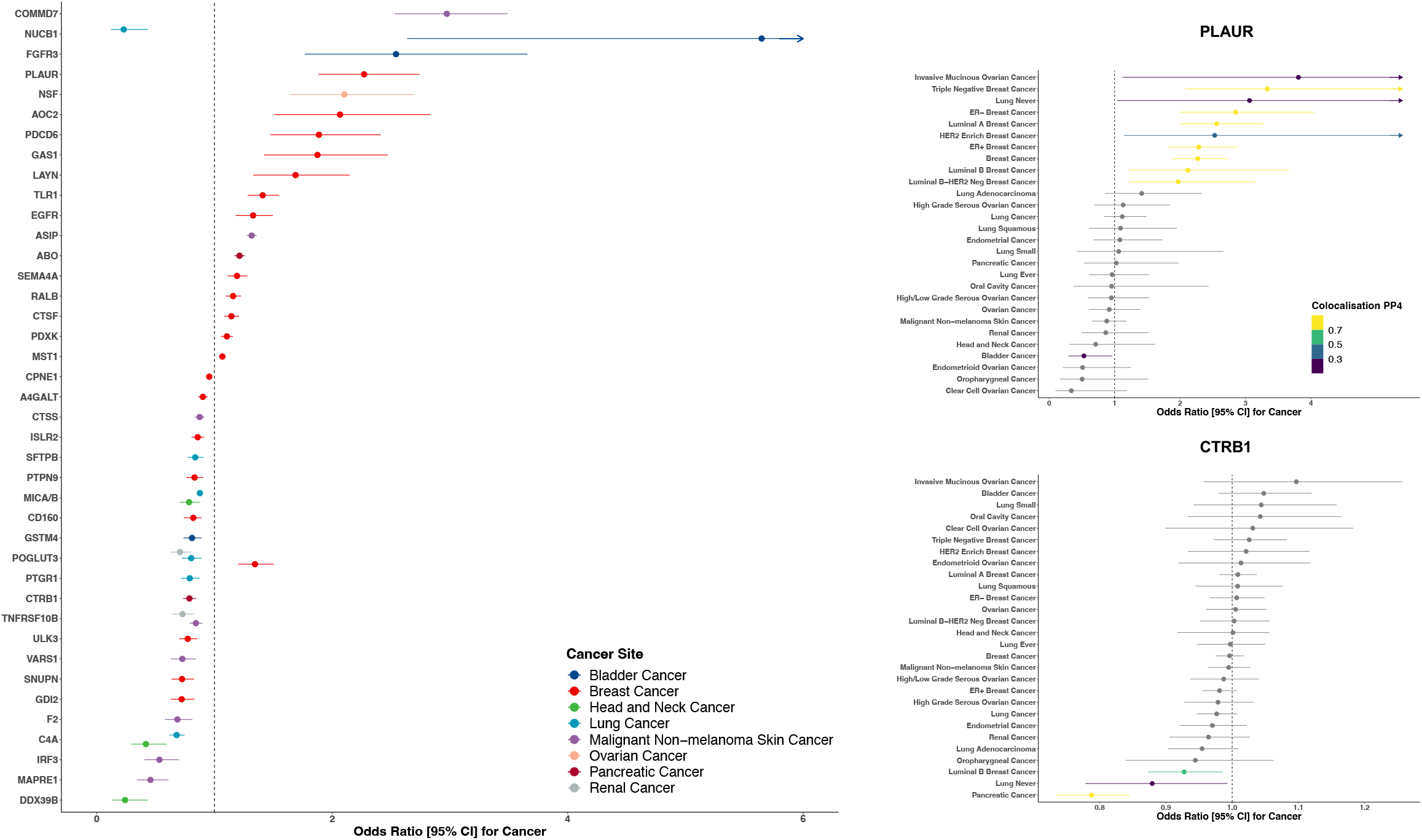
Proteins associated with cancer risk (left). Odds ratio estimates are scaled per standard deviation increment in relative circulating protein concentrations. Association for higher CTRB1 with cancer risk presented coloured by the colocalization probability (PP4), where MR Wald p < 0.05, to demonstrate pancreas-specific association (right). Association of higher PLAUR with cancer risk (right) with colour scheme as described above.

### Hormone-related cancers

#### Breast

We found 21 proteins associated with breast cancer risk. Among these proteins, nine (AOC2, SPN1, CD160, RALB, GDI2, CPNE1, ULK3, CTSF, PLAUR) were colocalised and had at least nominal associations with multiple molecular subtypes of breast cancer. PLAUR was observed to have a strong positive association with risk for breast cancer overall [odds ratio per standard deviation increment (OR): 2.27, 95% CI: 1.88 to 2.74; PP4: 0.99] and all molecular subtypes except HER2 enriched tumours. A majority of the remaining eight proteins were associated with molecular subtypes characterised by oestrogen-receptor (ER) positive tumours, such as AOC2, which was associated with a higher risk of ER-positive [OR: 2.02, 95% CI: 1.48 to 2.75; PP4: 0.73], luminal A [OR: 2.07, 95% CI: 1.51 to 2.83; PP4: 0.99], and luminal B-HER2 negative tumours [OR: 2.54, 95% CI: 1.43 to 4.51; PP4: 0.99]. In contrast, RALB was associated with an increased risk of ER-negative [OR: 1.40, 95% CI: 1.20 to 1.64; PP4: 0.93] and HER2 enriched tumours [OR: 1.59, 95% CI: 1.14 to 2.25; PP4: 0.98], as well as breast cancer overall [OR: 1.16, 95% CI: 1.09 to 1.23; PP4: 0.70]. GAS1 appeared to have a relatively specific association with risk of triple negative breast cancer [OR: 1.88, 95% CI: 1.42 to 2.47; PP4: 0.86] but was also associated with a lower risk of HER2 enriched breast cancer [OR: 0.46, 95% CI: 0.27 to 0.78; PP4: 0.78]. MST1 [OR: 1.07, 95% CI: 1.04 to 1.09; PP4: 0.94] was only associated with ER-negative tumours. Four of the 21 proteins (PDCD6, TLR1, POGLUT3, and LAYN) identified due to their association with breast cancer were also observed to have colocalised and at least nominal associations with risk for other cancer sites included in this study. Among these, two also associated at nominal significance with ovarian cancer (TLR1 and PDCD6), one also associated with lung cancer (POGLUT3) and one associated with each of kidney (POGLUT3), and bladder cancer (LAYN). ***Ovary***. One protein had an association with risk of ovarian cancer. NSF was associated with multiple ovarian cancer endpoints, including risk of high-grade serous tumours [OR: 2.26, 95% CI: 1.68 to 3.03; PP4: 0.99].

### Upper gastrointestinal and respiratory cancers

#### Lung

We found six proteins (PTGR1, C4A, MICA/B, SFTPB, NUCB1, POGLUT3) with strong evidence for an association with lung cancer risk of which two, SFTPB and PTGR1, were not observed to associate with risk of other cancers. SFTPB was associated with lower risk of lung cancer overall [OR: 0.79, 95% CI: 0.69 to 0.91; PP4: 0.82], but also lung adenocarcinoma, and lung cancer in never-smokers. In contrast, PTGR1 was associated with a lower risk of ever smoking and with squamous cell tumours [OR: 0.79, 95% CI: 0.72 to 0.87; PP4: 0.99]. C4A was inversely associated with both risk of lung cancer overall [OR: 0.68, 95% CI: 0.62 to 0.75; PP4: 0.96] and oral cavity cancer [OR: 0.42, 95% CI: 0.29 to 0.59; PP4: 0.99]. ***Head and Neck***. C4A, DDX39B, and MICA/B had associations with risk of head and neck cancers of which one, DDX39B, appeared cancer-subtype-specific and was associated with a lower risk of oropharyngeal cancer [OR: 0.24, 95% CI: 0.13 to 0.43; PP4: 0.94]. We also identified an inverse association of MICA/B with a lower risk of oral cavity cancer [OR: 0.79, 95% CI: 0.70 to 0.88; PP4: 0.99] and lung cancer overall [OR: 0.88, 95% CI: 0.85 to 0.90; PP4: 0.96]. Conversely this protein was associated with a modest, nominally significant, increased risk of endometrial cancer [OR: 1.10, 95% CI: 1.05 to 1.15; PP4: 0.89].

### Urological cancers

#### Kidney

TNFRSF10B and POGLUT3 had noteworthy associations with kidney cancer. Both proteins were associated with other malignancies. TNFRSF10B was associated with a lower risk of both kidney cancer [OR: 0.73, 95% CI: 0.64 to 0.83; PP4: 0.99] and non-malignant melanoma. POGLUT3 was associated with a lower risk of kidney cancer [OR: 0.71, 95% CI: 0.63 to 0.79; PP4: 0.98] and lung adenocarcinoma [OR: 0.80, 95% CI: 0.73 to 0.89; PP4: 0.99] but an increased risk of both ER- and triple negative breast cancer.

#### Bladder

NUCB1, GSTM4, and FGFR3 had associations with bladder cancer risk. One of these, GSTM4, was associated with a cancer-specific and lower risk [OR: 0.81, 95% CI: 0.74 to 0.89; PP4: 0.99]. Two others, NUCB1 [OR: 5.65, 95% CI: 2.64 to 12.09; PP4: 0.98] and FGFR3 [OR: 2.54, 95% CI: 1.77 to 3.66; PP4: 0.99] were both associated with a higher risk of bladder cancer and were observed to have associations with other cancers, including luminal B breast cancer and lung adenocarcinoma.

### Skin and Pancreas cancers

#### Non-malignant melanoma

Eight proteins (TNFRSF10B, F2, CTSS, VARS1, ASIP, IRF3, MAPRE1, COMMD7) had an association of note with risk of non-malignant melanoma of which four (F2, VARS1, IRF3, and MAPRE1) were not observed to associate with other cancers in this study, such as IRF3 [OR: 0.53, 95% CI: 0.41 to 0.70; PP4: 0.87]. Of the four other proteins, CTSS [OR: 0.87, 95% CI: 0.83 to 0.92; PP4: 0.98] and COMMD7 [OR: 2.97, 95% CI: 2.53 to 3.49; PP4: 0.99] were also associated with lung cancer overall [OR: 0.93, 95% CI: 0.89 to 0.98; PP4: 0.99] and never smoking lung cancer [OR: 2.78, 95% CI: 1.36 to 5.69; PP4: 0.78], respectively. **Pancreas**. CTRB1 and ABO were associated with risk of pancreatic cancer of which one was only associated with pancreas cancer and largely only expressed in the exocrine pancreas, CTRB1 [OR: 0.79, 95% CI: 0.73 to 0.85; PP4: 0.99]. ABO was associated with an increased risk of pancreas [OR: 1.21, 95% CI: 1.17 to 1.26; PP4: 0.97] and endometrial cancers [OR: 1.05, 95% CI: 1.03 to 1.08; PP4: 0.97] but with a lower risk of lung cancer [OR: 0.98, 95% CI: 0.96 to 0.99; PP4: 0.74].

Thirty-eight of the 40 cancer risk proteins were identified only using *cis* pQTL for proteins measured on the Somalogic platform. Two proteins (TNFRSF10B and PLAUR) were identified only using *cis* pQTL from GWAS of proteins measured using the Olink platform.

### Replication of Candidate Aetiological Proteins for Cancer Risk

Replication analyses were conducted for associations where an external cancer GWAS was available. Replication of *cis* pQTL MR associations were observed for 29 of the 68 protein-cancer associations that we were able to investigate (Table 2). For example, primary analyses identified higher GSTM4 was associated with lower risk of bladder cancer [OR: 0.81, 95% CI: 0.74 to 0.89, PP4: 0.99] in main analyses, which was replicated in a combined bladder cancer GWAS from UK Biobank and Finngen cohorts [OR: 0.85, 95% CI: 0.77 to 0.93]. Additionally, replication in UKBB and Finngen was observed for other results, including PLAUR [breast cancer OR: 1.82, 95% CI: 1.37 to 2.42], POGLUT3 [kidney cancer OR: 0.72, 95% CI: 0.60 to 0.88], and CTRB1 [pancreas cancer OR: 0.83, 95% CI: 0.75 to 0.92]. No replication GWAS were available for oropharyngeal and high grade ovarian serous cancer, or luminal A or triple negative breast cancer.

### Protein association with other traits

#### Colocalization and MR PHEWAS

We identified associations for many proteins, found to associate with cancer risk, with non-cancer endpoints that may be informative for determining the specificity of their associations with cancer or the utility of any potential therapeutic intervention. Notably, however, we did not observe any associations with other traits for SFTPB, EGFR, and GAS1, which were associated with lower risk of lung adenocarcinoma and higher risk of breast cancer overall and triple-negative breast cancer, respectively. All results are presented in Supplementary table 3, while two proteins associated with an increased cancer risk are discussed here in greater detail below as emblematic vignettes for the potential consequences of intervening to lower a protein to reduce cancer risk (Fig 3.).

**Figure 3.**
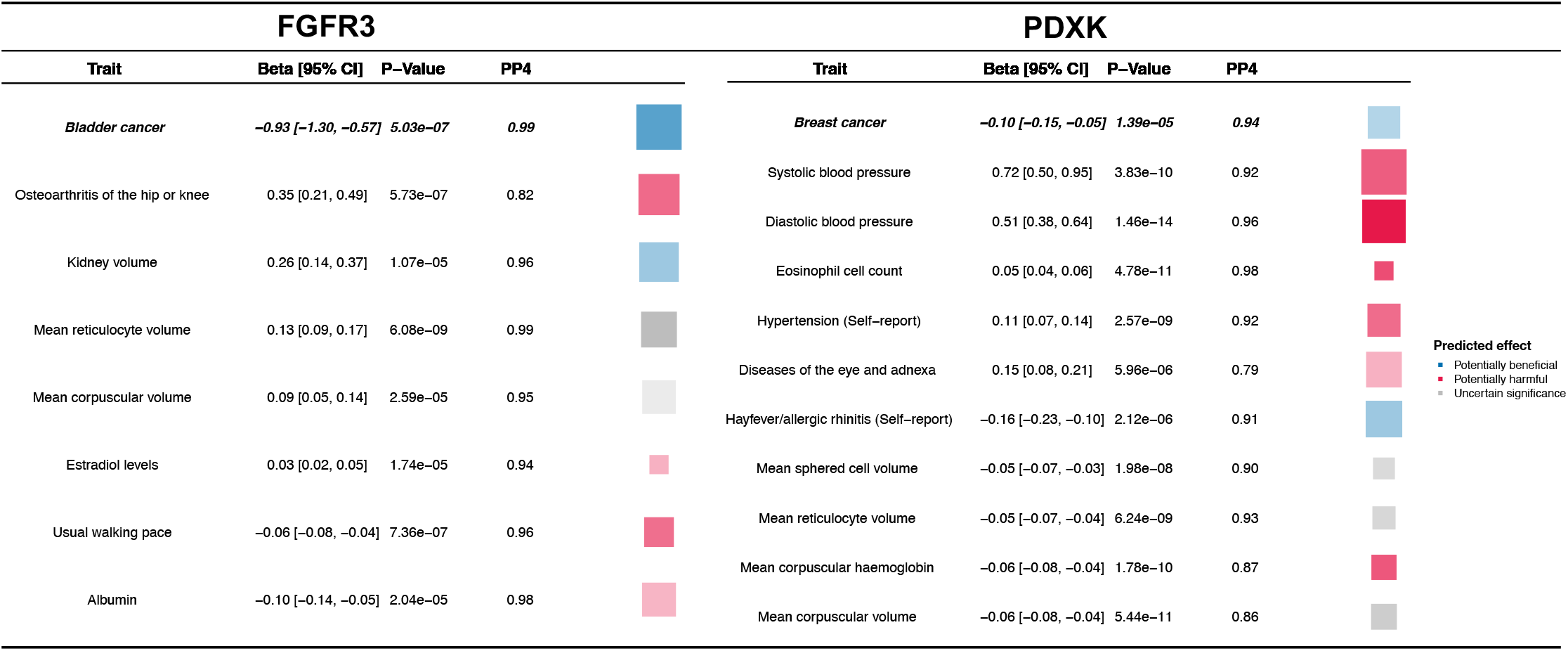
Potential consequences of a protein-lowering intervention for two emblematic cancer risk proteins, FGFR3 and PDXK, that associate with higher risk of bladder and breast cancer, respectively. For reference, the predicted effect of lowering each protein by 1 SD is present for cancer risk at the top (italics and bold). Below these estimates we present the predicted effect of protein-lowering where colocalization and MR analyses suggest an aetiological link on other traits. Box colour indicates whether a predicted consequence may be beneficial (blue), harmful (red) or have uncertain consequence (grey) on health. Box size is proportionate to absolute beta from MR analyses while box opacity is proportionate to precision of this MR estimate.

Higher FGFR3 was associated with an increased risk of bladder cancer. However, we observe that lowering FGFR3 may have potentially harmful effects on other common sources of morbidity, such as a higher risk of osteoarthritis of the hip or knee [OR: 1.42, 95% CI: 1.24 to 1.63; PP4: 0.99] and a reduced usual walking pace [Beta: -0.05, 95% CI: -0.03 to -0.07; PP4: 0.96], and higher circulating oestradiol levels (SD) [Beta: 0.03, 95% CI: 0.01 to 0.05; PP4: 0.94] and lower circulating albumin (SD) [Beta: -0.09, 95% CI: -0.06 to -0.14; PP4: 0.98]. Nonetheless, lower FGFR3 may also lead to other potentially beneficial consequences including a higher kidney volume (litres) [Beta: 0.26, 95% CI: 0.14 to 0.38; PP4: 0.96].

Higher PDXK was associated with an increased risk of breast cancer. However, lowering PDXK was also associated with higher systolic [Beta: 0.72, 95% CI: 0.49 to 0.95; PP4: 0.92] and diastolic blood pressure (mmHg) [Beta: 0.51, 95% CI: 0.38 to 0.64; PP4: 0.96], a higher risk of having hypertension [OR: 1.12, 95% CI: 1.07 to 1.15; PP4: 0.90]. It was also associated with higher eosinophil counts (SD) [Beta: 0.05, 95% CI: 0.03 to 0.08; PP4: 0.98] and diseases of the eye and adnexa [OR: 1.16, 95% CI: 1.08 to 1.23; PP4: 0.79], but a lower risk of reporting hay fever or allergic rhinitis [OR: 0.85, 95% CI: 0.79 to 0.91; PP4: 0.85].

#### HyprColoc Analyses

Eight proteins (A4GALT, ASIP, CTSF, MARE1, PDXK, SEM4A, PLAUR, VARS1) were observed to have evidence of multi-trait colocalization between the index protein, intermediate phenotypes, and the index cancer endpoint (Supplementary Table S4), which may serve to elucidate aetiological pathways to cancer risk. For example, higher PLAUR, a breast cancer risk protein, had evidence of a colocalized association with lower monocyte cell count [Beta: -0.52; PP4: 0.99] and higher granulocyte percentage of myeloid white cells [Beta: 0.63; PP4: 0.99], and strong evidence for a shared association of PLAUR and these blood cell traits with risk of breast cancer overall [PP4: 0.91], triple negative [PP4: 0.89], luminal A [PP4: 0.90], and ER negative breast cancer [PP4: 0.89].

#### Public Databases

Thirty-one of our cancer risk proteins had pLI scores of < 0.1 implying high tolerance for loss of function variation (Table 2). Conversely, two proteins, DDX39B and MAPRE1, appeared highly intolerant of LoF variation and had pLI > 0.9. We observed only limited evidence for the association of pLOF variants in cognate genes for cancer risk proteins with other traits in the UK Biobank using Genebass or the AstraZeneca PheWAS Portal, none of which were cancer endpoints (LAYN, PTGR1, FGFR3, VARS1, PTPN9), none of which were cancer endpoints. In Genebass, pLOF in LAYN was associated with lower forced expiratory volume in 1-second (beta: -2.99^-4^, *p* = 1.91^-11^), while variation in PLAUR was associated with lower carotid intima-medial thickness (beta: -1.09^-1^, *p*= 2.03^-6^). PTGR1 pLOF burden was associated with high monocyte percentage (beta: 5.58^-3^, *p* = 1.34^-7^), PTPN9 pLOF burden was associated with recent changes in the speed/amount of moving or speaking (OR: 1.02, *p* = 6.84^-15^), and VARS1 with lower mean corpuscular haemoglobin (beta: -1.09^-2^, *p* = 2.41^-6^). Additionally, rare protein-damaging missense variation in FGFR3 was the top predictor of osteochondrodysplasia (OR: 61.51, 7.13^-12^) from analyses in the UK Biobank reported in the AstraZeneca PheWAS Portal.

We also identified evidence of Mendelian disorders associated with genetic alterations to the cognate genes of 17 of the 40 noteworthy proteins, such as FGFR3 and F2 in OMIM. Among other conditions, such as stroke and pregnancy loss, F2 mutations have been associated with higher circulating prothrombin levels and an increased risk of venous thrombosis. Mutations in FGFR3 have been associated with achondroplasia.

### Drug Target pQTL Analyses

Harmonised *cis* pQTL were available for up to 473 of the total 488 proteins whose cognate gene was mapped to a known drug target for at least one cancer outcome. After correction for multiple testing, we found 18 proteins mapped to drugs that associated with risk of at least one cancer endpoints. These included breast [8 proteins], lung [3 proteins], head and neck [2 proteins], kidney [2 proteins], malignant non-melanoma [3 protein], and bladder [1 protein], and pancreas [1 protein] cancers that were the target of at least one pharmaceutical intervention (Supplementary Table S5). We additionally identified eight proteins that were the target of a drug under investigation at phase I clinical trials or higher, while eight out of these proteins are currently at preclinical stage or biological testing, which may imply their drug-ability is under active evaluation. Fifteen of the proteins we identify associated with cancer risk, including SFTPB and GAS1, did not appear to be under active current investigation as a drug target (Fig 4.).

**Figure 4.**
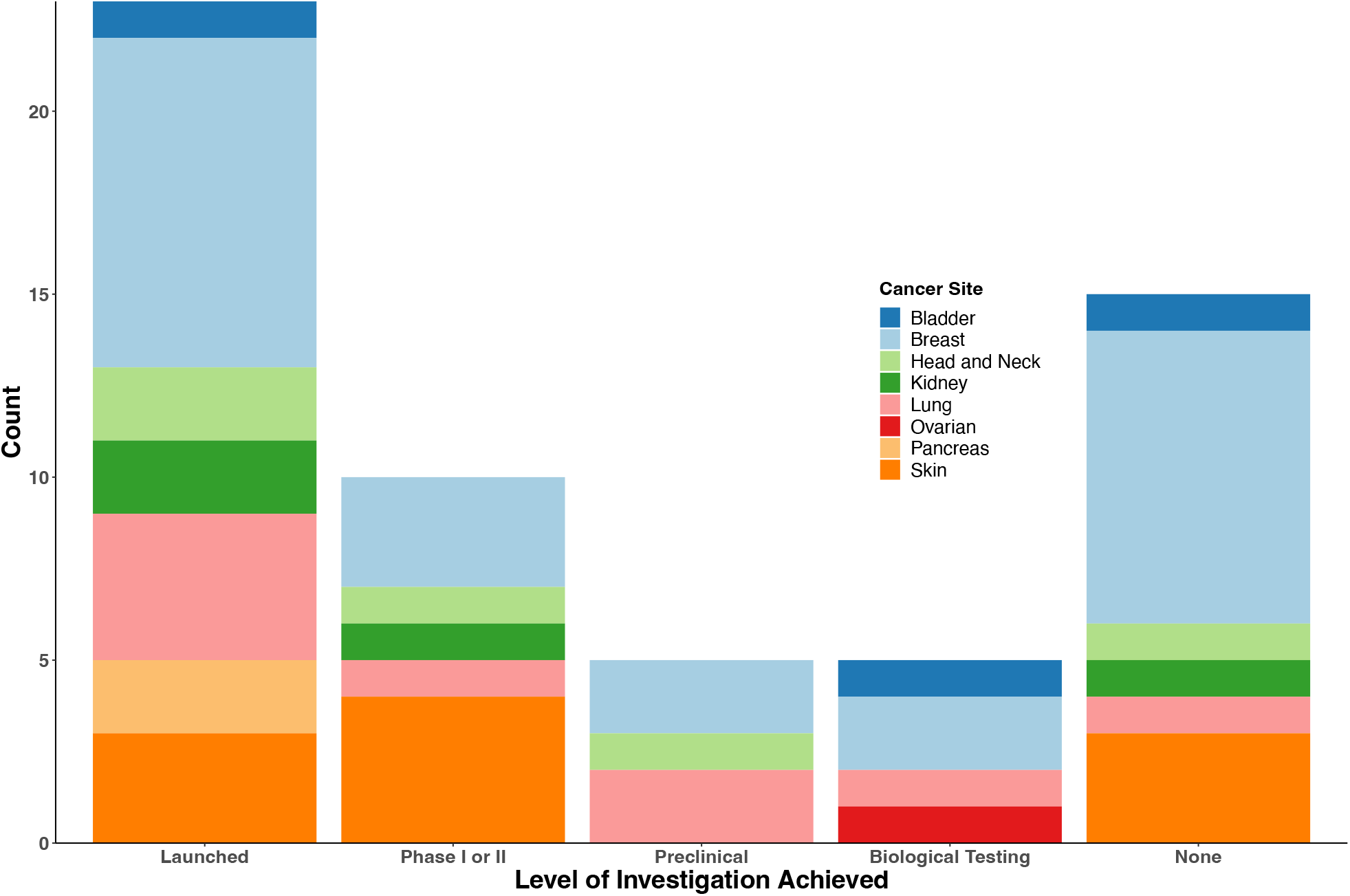
Stacked bar plot that depicts identified proteins associated with cancer by their current highest level of therapeutic investigation, which additionally has colours that stratified results by cancer site.

Notably, a majority of drugs targeting cancer risk proteins were typically either small molecular inhibitors (SMIs) or monoclonal antibodies, some of which are used for the treatment of the cancer indicated from the risk association. For example, higher FGFR3 was associated with an increased risk of bladder cancer. FGFR3 is directly inhibited by Erdafitinib^36^, which is a SMI and a treatment of urothelial cancers. We also identified an association of EGFR with a higher risk of breast cancer, which is inhibited by a variety of monoclonal antibodies in the treatment of breast cancer^37^.

## Discussion

Using genetic data from up to 337,822 cancer cases, we have identified 40 proteins with a likely role in the aetiology of at least one type of cancer. Most proteins that we identify associated with cancer risk were replicated using an independent cancer GWAS and had not been reported on before in this context. Some risk proteins were associated with a potential causal role in specific molecular subtypes of cancer, some were risk proteins with specific expression in the relevant organ, while others associated with risk of multiple cancers. We also identified proteins with a potential aetiological role in cancer risk that also mediate the therapeutic effects of specific drugs that may lead to possible drug repurposing. Furthermore, we identified proteins that are associated with cancer but do not appear to be under active investigation, indicating that they may represent novel opportunities to develop new therapeutic treatments of cancer.

Proteins are crucial for the maintenance of cellular structure and regulation of cell signalling involved in all human biological processes. Identifying protein markers of cancer aetiology may inform our understanding of the pathways to tumorigenesis and serve as a fruitful tool for discovering novel biomarkers of cancer risk. As a proof-of-concept for how genetic methods may identify causal cancer genes we used this approach to highlight a causal role of EGFR in breast cancer risk. EGFR is a COSMIC^38^ consensus gene associated with breast cancer risk. Among other processes, EGFR has an established role in cell proliferation, migration and differentiation that facilitates the uncontrolled division of cancer epithelial cells^39^, including those in the mammary glands^37^. EGFR may therefore serve as a potential positive control for protein MR as a method to discover oncogenic pathways.

We also identified a role for proteins with tissue-specific expression at the site of the cancer indicated in risk analyses. One example is CTRB1, which is a serine protease digestive enzyme precursor produced largely by the exocrine pancreas and which was associated with a lower risk of pancreatic cancer in two independent cancer GWAS. Along with PRSS2 and SPINK, CTRB1 acts to degrade trypsinogen in the pancreas. Mutations in CTRB1 have been associated with higher intrapancreatic trypsin activation, chronic inflammation, and loss of pancreas function^40^. Further, CTRB1 expression has been found in the topmost downregulated genes in matched tumour-normal pancreas samples^41^. However, the CTRB1 region on chromosome 16 contains various complex genomic rearrangements that include a short deletion in CTRB2 associated with pancreas cancer risk^42^ and a 16.6kb inversion between CTRB1 and CTRB2, which is associated with chronic pancreatitis^43^, a risk factor for pancreatic cancer. While we do not find an association of CTRB2 with pancreatic cancer risk, we do add to previous evidence^44^ that CTRB1 may affect the risk of both type 1 and type 2 diabetes in PHEWAS analyses, which may be risk factors for pancreas cancer. Therefore, further analyses are needed to clarify the role of the CTRB1-CTRB2 locus in pancreatic cancer risk, which could include pre-clinical assessment of CTRB1 and CTRB2 as potential anti-tumour agents as was previously conducted for pro-enzymes PRSS1 and Chymotrypsinogen A^45^.

Notably, among cancer risk proteins we observe a high tolerance for haploinsufficiency and a general absence of congruent evidence using whole exome analyses of potential loss of function variants in the UK Biobank for the role of proteins we identify with a risk of cancer. We suspect this may at least in part be due to study power for whole exome analyses and challenges in the interpretation of pLI scores in the context of cancer^46^. However, it may also suggest these proteins affect cancer risk via regulation of abundance as compared to the presence or absence of functional gene copies. Moreover, this finding may imply protein-truncating alterations to these proteins’ cognate genes are unlikely to lead to severe disease in early life, which may affect their relevance in therapeutic prevention.

Proteins are essential targets for drug development^47^. However, not all proteins identified as risk factors for cancer are suitable therapeutic targets. Biochemical factors that contribute to a protein’s *druggability* include protein-class (membrane-bound^36,48^, or soluble and secreted ligands^49^, for example), current understanding of its pathophysiology and molecular function, its cellular location, and the presence of drug binding sites. Epidemiological factors that contribute to a protein’s utility as a therapeutic target include: the magnitude and specificity of its association with cancer risk, the expected incidence, and morbidity and mortality of the cancer of interest; and our current ability to robustly identify an *at-risk* population to receive a proposed therapeutic intervention. We present three vignettes to illustrate both the potential and the complexities when investigating proteins for cancer prevention.

### FGFR3 and Bladder Cancer

We found an increase in FGFR3 associated with a more than two-fold risk of bladder cancer in two independent cancer GWAS. FGFR3 is an established oncogene with a well-described role in proliferative signalling and evasion of cell death^39^. Further, FGFR3 is the target of an approved tyrosine kinase inhibitor used to treat urothelial cancer, erdafitinib^50^, which has modest reported toxicity^51^. Whereas these data may suggest FGFR3 as a potential target for therapeutic prevention, further complementary analyses suggested lower FGFR3 may increase risk of osteoarthritis, a common source of morbidity among older populations, and rare variant studies show that damaged FGFR3 increases the risk of bone disorders. Considering bladder cancer is not among the most common cancers and has relatively good prognosis, it may therefore not be justified to target FGFR3 specifically to prevent bladder cancer. This example highlights the importance of considering a broad spectrum of potential health outcomes when developing therapeutic prevention strategies for specific cancer sites to identify potential unintentional but harmful secondary effects.

### SFTPB and Lung Cancer

We found that higher SFTPB was associated with a lower risk of lung adenocarcinoma. SFTPB is produced specifically by alveolar type II cells^52^ and is essential for healthy lung function^53,54^. Previous studies identified downregulation of SFTPB in mice to be associated with accelerated tumour growth and rate of epithelial-mesenchymal transition^55^, and higher SFTPB expression in tumour samples is associated with better survival among lung cancer patients^54^. Notably, we did not find evidence of non-lung cancer associations for SFTPB, though an association with chronic obstructive pulmonary disease has previously been reported^56^. Lung cancer is a highly incident cancer and the leading cause of cancer death globally. Multiple tools for the identification and referral of individuals at high risk of lung cancer for screening programmes are in current use with additional biomarker-based risk models under development, such as within the INTEGRAL programme^57^. Lung cancer may therefore present an appealing target for therapeutic prevention as high-risk individuals who may benefit from such interventions are readily identifiable if a suitable agonist for SFTPB can be identified.

### GAS1 and Triple Negative Breast Cancer

GAS1, which was associated with an increased risk of triple negative breast cancer (TNBC), is an essential co-receptor of hedgehog (Hh) signalling and specifically expressed at high levels in healthy fibroblasts ^58,59^. Notably, we did not observe evidence that GAS1 had a similar effect on risk of other molecular subtypes of breast cancer or on non-cancer phenotypes, which may imply GAS1 has a specific effect to increase TNBC risk. GAS1 has a critical role, via a structural interaction, in the delivery of sonic hedgehog to its downstream receptor, PTCH1^60^. Furthermore, Hh-activated TNBC mouse models indicated a specific ligand-dependent paracrine role for the Hh pathway acting via cancer-associated fibroblasts (CAF) to maintain chemotherapy-resistant breast cancer stem cell phenotypes. An independent study also identified a paracrine Hh pathway signature was associated with higher risk of metastasis and breast cancer-specific death in triple-negative disease^61^. TNBC is an aggressive molecular subtype of breast cancer characterised by resistance to many established treatments^62^. Therefore, compared to currently available drugs that target the SMO gene in any cell type, GAS1 antagonism could provide a novel therapeutic approach for CAF-specific Hh pathway inhibition in chemoresistant TNBC^63^.

Our study has limitations that should be acknowledged. Firstly, our study was not comprehensive in evaluating the entire proteome as we are limited to proteins that have been measured using blood based multiplex affinity platforms. Secondly, where we have *cis* pQTL, our power to discover associations may be limited by cancer GWAS sample size and the heritability of the cancer itself. This may partly explain the discrepancy in the number of risk proteins identified for different cancer sites and the ability to replicate protein associations with risk. We also note that colocalization can be sensitive to the presence of complex genetic architecture or loci where multiple causal signals exists but where only a subset are shared between traits. Similarly, it can be difficult to interpret colocalization between a protein and cancer risk where a cognate gene sits in a genomic region with irregular haplotype structure, such as C4A associations with lung cancer on chromosome six, or when two genes that lie immediately adjacent to each other, both share similar associations with cancer risk, such as PTPN9 and SNUPN with breast cancer risk in our analyses. We also note that our replication analyses leveraged data from cancer GWAS in a Finnish population, and thus, that well-documented changes in allele frequencies due to founder effects may have hindered successful replication for some proteins. Finally, we note that our results are restricted to participants of European ancestry due principally to the current availability of protein GWAS; while we are aware of a recent protein GWAS in the ARIC cohort^64^ in people of African American descent, we did not have access to large cancer GWAS for endpoints in this study to utilise these data.

Conversely, our study had several notable strengths. Our widespread integration of colocalization with *cis* pQTL MR analyses demonstrated that upwards of 70% of proteins with a nominally significant MR association did not have support for a shared causal locus. While this is closer to 50% for proteins passing our Bonferroni threshold, we suggest that these results demonstrate the importance of presenting colocalization in parallel with any *cis* pQTL MR analysis. The inclusion of multiple cancer endpoints in our study has also allowed us to identify proteins with both associations across multiple cancer endpoints and those with cancer-specific associations. Similarly, we conducted analyses and integrated multiple sources of clinically relevant data to better contextualise which of the 40 risk proteins are associated with other non-cancerous traits. Additionally, we mapped our results to established drug targets as well as proteins currently undergoing investigation as novel therapeutics. In doing so, we highlight opportunities for drug repurposing, but note that risk proteins that are not currently targeted by any drug may serve as appealing targets for future drug development.

The increasing availability of multi-omic data generated in large biobanks and cancer consortia presents an unprecedented opportunity to leverage large-scale genetic data to discover novel disease pathways. We present an expansive assessment of 2,094 proteins in relation to nine common cancer sites, and present multiple novel proteins implicated in the aetiology of specific cancer types. We demonstrate the importance of carrying out complementary analyses to characterise the disease specificity and pleiotropy that is present for many proteins. We believe these results bring important context both when understanding aetiological pathways and potential adverse consequences of any potential protein-altering intervention.

## Data Availability

All data produced in the present work are contained in the manuscript

## References

1. Ferlay J et al. Global Cancer Observatory: Cancer Tomorrow. International Agency for Research on Cancer. https://gco.iarc.fr/tomorrow, accessed [28 02 2023].

2. Cheung, B. M. Y. & Lam, K. S. L. Never too old for statin treatment? The Lancet 393, 379–380 (2019).

3. Reiner, Ž. Statins in the primary prevention of cardiovascular disease. Nat Rev Cardiol 10, 453–464 (2013).

4. Omenn, G. S. et al. Effects of a Combination of Beta Carotene and Vitamin A on Lung Cancer and Cardiovascular Disease. New England Journal of Medicine 334, 1150–1155 (1996).

5. Harbeck, N. et al. Breast cancer. Nat Rev Dis Primers 5, 66 (2019).

6. Burn, J. et al. Cancer prevention with aspirin in hereditary colorectal cancer (Lynch syndrome), 10-year follow-up and registry-based 20-year data in the CAPP2 study: a double-blind, randomised, placebo-controlled trial. The Lancet 395, 1855–1863 (2020).

7. Murphy, N. et al. Insulin-like growth factor-1, insulin-like growth factor-binding protein-3, and breast cancer risk: observational and Mendelian randomization analyses with ∼430 000 women. Annals of Oncology 31, 641–649 (2020).

8. Murphy, N. et al. Circulating Levels of Insulin-like Growth Factor 1 and Insulin-like Growth Factor Binding Protein 3 Associate With Risk of Colorectal Cancer Based on Serologic and Mendelian Randomization Analyses. Gastroenterology 158, 1300–1312.e20 (2020).

9. Travis, R. C. et al. A Meta-analysis of Individual Participant Data Reveals an Association between Circulating Levels of IGF-I and Prostate Cancer Risk. Cancer Res 76, 2288–2300 (2016).

10. Watts, E. L. et al. Circulating insulin-like growth factor-I, total and free testosterone concentrations and prostate cancer risk in 200 000 men in UK Biobank. Int J Cancer 148, 2274–2288 (2021).

11. Smith Byrne, K. et al. The role of plasma microseminoprotein-beta in prostate cancer: an observational nested case–control and Mendelian randomization study in the European prospective investigation into cancer and nutrition. Annals of Oncology 30, 983–989 (2019).

12. Brenner, D. R. et al. Inflammatory Cytokines and Lung Cancer Risk in 3 Prospective Studies. Am J Epidemiol 185, 86–95 (2017).

13. Watts, E. L. et al. Circulating free testosterone and risk of aggressive prostate cancer: Prospective and Mendelian randomisation analyses in international consortia. Int J Cancer 151, 1033–1046 (2022).

14. Smith-Byrne, K. et al. Circulating Isovalerylcarnitine and Lung Cancer Risk: Evidence from Mendelian Randomization and Prediagnostic Blood Measurements. Cancer Epidemiology, Biomarkers & Prevention 31, 1966–1974 (2022).

15. Pietzner, M. et al. Mapping the proteo-genomic convergence of human diseases. Science (1979) 374, (2021).

16. Swerdlow, D. I. et al. Selecting instruments for Mendelian randomization in the wake of genome-wide association studies. Int J Epidemiol 45, 1600–1616 (2016).

17. Fauman, E. B. & Hyde, C. An optimal variant to gene distance window derived from an empirical definition of cis and trans protein QTLs. BMC Bioinformatics 23, 169 (2022).

18. Wallace, C. A more accurate method for colocalisation analysis allowing for multiple causal variants. PLoS Genet 17, e1009440 (2021).

19. Zheng, J. et al. Phenome-wide Mendelian randomization mapping the influence of the plasma proteome on complex diseases. Nat Genet 52, 1122–1131 (2020).

20. Ferkingstad, E. et al. Large-scale integration of the plasma proteome with genetics and disease. Nat Genet 53, 1712–1721 (2021).

21. Folkersen, L. et al. Genomic and drug target evaluation of 90 cardiovascular proteins in 30,931 individuals. Nat Metab 2, 1135–1148 (2020).

22. Rothman, N. et al. A multi-stage genome-wide association study of bladder cancer identifies multiple susceptibility loci. Nat Genet 42, 978–984 (2010).

23. Zhang, H. et al. Genome-wide association study identifies 32 novel breast cancer susceptibility loci from overall and subtype-specific analyses. Nat Genet 52, 572–581 (2020).

24. O’Mara, T. A. et al. Identification of nine new susceptibility loci for endometrial cancer. Nat Commun 9, 3166 (2018).

25. Lesseur, C. et al. Genome-wide association meta-analysis identifies pleiotropic risk loci for aerodigestive squamous cell cancers. PLoS Genet 17, e1009254 (2021).

26. McKay, J. D. et al. Large-scale association analysis identifies new lung cancer susceptibility loci and heterogeneity in genetic susceptibility across histological subtypes. Nat Genet 49, 1126–1132 (2017).

27. Phelan, C. M. et al. Identification of 12 new susceptibility loci for different histotypes of epithelial ovarian cancer. Nat Genet 49, 680–691 (2017).

28. Klein, A. P. et al. Genome-wide meta-analysis identifies five new susceptibility loci for pancreatic cancer. Nat Commun 9, 556 (2018).

29. Scelo, G. et al. Genome-wide association study identifies multiple risk loci for renal cell carcinoma. Nat Commun 8, 15724 (2017).

30. Kimberley Burrows & Philip Haycock. Genome-wide Association Study of Cancer Risk in UK Biobank. 10.5523/bris.aed0u12w0ede20olb0m77p4b9.

31. Giambartolomei, C. et al. Bayesian Test for Colocalisation between Pairs of Genetic Association Studies Using Summary Statistics. PLoS Genet 10, e1004383 (2014).

32. Kurki, M. I. et al. FinnGen provides genetic insights from a well-phenotyped isolated population. Nature 613, 508–518 (2023).

33. Hewitt, J., Walters, M., Padmanabhan, S. & Dawson, J. Cohort profile of the UK Biobank: diagnosis and characteristics of cerebrovascular disease. BMJ Open 6, e009161 (2016).

34. Hemani, G. et al. The MR-Base platform supports systematic causal inference across the human phenome. Elife 7, (2018).

35. Foley, C. N. et al. A fast and efficient colocalization algorithm for identifying shared genetic risk factors across multiple traits. Nat Commun 12, 764 (2021).

36. Attwood, M. M., Fabbro, D., Sokolov, A. V, Knapp, S. & Schiöth, H. B. Trends in kinase drug discovery: targets, indications and inhibitor design. Nat Rev Drug Discov 20, 839–861 (2021).

37. Mukhopadhyay, C., Zhao, X., Maroni, D., Band, V. & Naramura, M. Distinct Effects of EGFR Ligands on Human Mammary Epithelial Cell Differentiation. PLoS One 8, e75907 (2013).

38. Tate, J. G. et al. COSMIC: the Catalogue Of Somatic Mutations In Cancer. Nucleic Acids Res 47, D941–D947 (2019).

39. Futreal, P. A. et al. A census of human cancer genes. Nat Rev Cancer 4, 177–183 (2004).

40. Jancsó, Z., Hegyi, E. & Sahin-Tóth, M. Chymotrypsin Reduces the Severity of Secretagogue-Induced Pancreatitis in Mice. Gastroenterology 155, 1017–1021 (2018).

41. Sakharkar, M. K., Dhillon, S. K., Mazumder, M. & Yang, J. Key drug-targeting genes in pancreatic ductal adenocarcinoma. Genes Cancer 12, 12–24 (2021).

42. Jermusyk, A. et al. A 584 bp deletion in CTRB2 inhibits chymotrypsin B2 activity and secretion and confers risk of pancreatic cancer. The American Journal of Human Genetics 108, 1852–1865 (2021).

43. Rosendahl, J. et al. Genome-wide association study identifies inversion in the CTRB1-CTRB2 locus to modify risk for alcoholic and non-alcoholic chronic pancreatitis. Gut 67, 1855–1863 (2018).

44. ‘t Hart, L. M. et al. The CTRB1/2 Locus Affects Diabetes Susceptibility and Treatment via the Incretin Pathway. Diabetes 62, 3275–3281 (2013).

45. Perán, M. et al. A formulation of pancreatic pro-enzymes provides potent anti-tumour efficacy: a pilot study focused on pancreatic and ovarian cancer. Sci Rep 7, 13998 (2017).

46. Ziegler, A., Colin, E., Goudenège, D. & Bonneau, D. A snapshot of some pLI score pitfalls. Hum Mutat humu.23763 (2019) doi:10.1002/humu.23763.

47. Bull, S. C. & Doig, A. J. Properties of Protein Drug Target Classes. PLoS One 10, e0117955 (2015).

48. Hauser, A. S., Attwood, M. M., Rask-Andersen, M., Schiöth, H. B. & Gloriam, D. E. Trends in GPCR drug discovery: new agents, targets and indications. Nat Rev Drug Discov 16, 829–842 (2017).

49. Attwood, M. M., Jonsson, J., Rask-Andersen, M. & Schiöth, H. B. Soluble ligands as drug targets. Nat Rev Drug Discov 19, 695–710 (2020).

50. Loriot, Y. et al. Erdafitinib in Locally Advanced or Metastatic Urothelial Carcinoma. New England Journal of Medicine 381, 338–348 (2019).

51. CENTER FOR DRUG EVALUATION AND RESEARCH. RISK ASSESSMENT and RISK MITIGATION REVIEW: Erdafitinib (212018Orig1s000). https://www.accessdata.fda.gov/drugsatfda_docs/nda/2019/212018Orig1s000RiskR.pdf [Accessed 28 02 2023].

52. Xiong, M., Heruth, D. P., Zhang, L. Q. & Ye, S. Q. Identification of lung-specific genes by meta-analysis of multiple tissue <scp>RNA</scp> -seq data. FEBS Open Bio 6, 774–781 (2016).

53. Nogee, L. M., deMello, D. E., Dehner, L. P. & Colten, H. R. Deficiency of Pulmonary Surfactant Protein B in Congenital Alveolar Proteinosis. New England Journal of Medicine 328, 406–410 (1993).

54. Clark, J. C. et al. Targeted disruption of the surfactant protein B gene disrupts surfactant homeostasis, causing respiratory failure in newborn mice. Proceedings of the National Academy of Sciences 92, 7794–7798 (1995).

55. Lee, S. et al. Surfactant Protein B Suppresses Lung Cancer Progression by Inhibiting Secretory Phospholipase A2 Activity and Arachidonic Acid Production. Cellular Physiology and Biochemistry 42, 1684–1700 (2017).

56. Yang, J. et al. Association of surfactant protein B gene with chronic obstructive pulmonary disease susceptibility. The International Journal of Tuberculosis and Lung Disease 18, 1378–1384 (2014).

57. Robbins, H. A. et al. Design and methodological considerations for biomarker discovery and validation in the Integrative Analysis of Lung Cancer Etiology and Risk (INTEGRAL) Program. Ann Epidemiol 77, 1–12 (2023).

58. Uhlén, M. et al. Tissue-based map of the human proteome. Science (1979) 347, (2015).

59. Lonsdale, J. et al. The Genotype-Tissue Expression (GTEx) project. Nat Genet 45, 580–585 (2013).

60. Huang, P. et al. Structural basis for catalyzed assembly of the Sonic hedgehog–Patched1 signaling complex. Dev Cell 57, 670–685.e8 (2022).

61. O’Toole, S. A. et al. Hedgehog Overexpression Is Associated with Stromal Interactions and Predicts for Poor Outcome in Breast Cancer. Cancer Res 71, 4002–4014 (2011).

62. Cazet, A. S. et al. Targeting stromal remodeling and cancer stem cell plasticity overcomes chemoresistance in triple negative breast cancer. Nat Commun 9, 2897 (2018).

63. Nguyen, N. M. & Cho, J. Hedgehog Pathway Inhibitors as Targeted Cancer Therapy and Strategies to Overcome Drug Resistance. Int J Mol Sci 23, 1733 (2022).

64. Wright, J. D. et al. The ARIC (Atherosclerosis Risk In Communities) Study. J Am Coll Cardiol 77, 2939–2959 (2021).

